# Risk of COVID-19 variant importation – How useful are travel control measures?

**DOI:** 10.1101/2021.05.13.21257141

**Authors:** Julien Arino, Pierre-Yves Boëlle, Evan Milliken, Stéphanie Portet

## Abstract

We consider models for the importation of a new variant COVID-19 strain in a location already seeing propagation of a resident variant. By distinguishing contaminations generated by imported cases from those originating in the community, we are able to evaluate the contribution of importations to the dynamics of the disease in a community. We find that after an initial seeding, the role of importations becomes marginal compared to that of community-based propagation. We also evaluate the role of two travel control measures, quarantine and travel interruptions. We conclude that quarantine is an efficacious way of lowering importation rates, while travel interruptions have the potential to delay the consequences of importations but need to be applied within a very tight time window following the initial emergence of the variant.

## 1. Introduction

SARS-CoV-2 is an RNA virus and as a consequence is prone to mutations. However, mutations in coronaviruses occur not as fast as for viruses such as HIV or Influenza because of copy editing [25]. During most of its early spread in 2020, few mutations were observed that became established. Then, in December 2020, the United Kingdom announced that its intensive sequencing efforts had led to the discovery of a variant, B.1.1.7, which seemed to be both in the process of establishing itself there and to be more transmissible than the “historic” variant. Since then, this variant has become the dominant source of infection in the UK and, at the time of writing, in several European countries and elsewhere. Other variants have also been detected. See a review in [25].

In reaction, many countries took measures to limit the entry of passengers hailing from countries where the novel variants were prevalent. Quarantine measures were also either implemented *de novo* or continued. See an extensive review of the types of measures used in [12].

The pattern has repeated itself since with, using the US CDC classification [32], various variants of interest (VOI), of concern (VOC) or of high consequence (VOHC) such as B.1.1.7, B.1.351, P.1 or B.1.617 and can be described as follows. Following the identification of a novel variant in some jurisdiction, local public health authorities communicate this information to their international counterparts and to the media. An amplification phase in the media and social media follows that results almost invariably, in other jurisdictions, to calls for stringent measures to be imposed, from quarantine all the way to the closing of borders. Whether or not countries followed through on these calls has varied greatly and has resulted from arbitration between arguments in favour of and arguments against such measures.

There are factual and scientific arguments in favour of strict travel control, besides the implicit need for politicians to heed the call of social and established media. From the perspective of international law, the World Health Organisation provides, through the International Health Regulations (IHR), a practical framework to deal with Public Health Emergencies of International Concern (PHEIC) such as COVID-19. The IHR provide guidelines for the inter-state dissemination of information about PHEIC, but also delineate the actions that states can take to safeguard the health security of their populations. In the context of international mobility, they set for instance the rights and responsibilities of states vis-‘a-vis international non-citizen travellers arriving on their soil in terms of testing, quarantine or treatment. At the start of the first wave, stringent border control measures allowed to identify cases. For instance, among the first cases detected in Hong Kong, numbers #1 and #7 were imported cases detected at the border [19]. Some modelling work also argues in favour of travel bans; see, e.g., [16, 27].

However, there are also arguments against strict travel control. Firstly, in the context of international law, the pendant for individuals to the IHR is framed by the mobility articles (Articles 13-15) in the Universal Declaration of Human Rights (UDHR) and its derived international and national treaties of application (e.g., the Canadian Charter of Rights and Freedoms). There have been discussions, in particular, about the necessity of allowing states to temporarily go counter to Article 13 (“Everyone has the right to leave any country, including his own, and to return to his country”) or its equivalent in national laws [10, 22, 28]. At this point, law courts have mostly sided with governments. However, a recent ruling in France shows that as the crisis continues, courts will likely start to rule in favour of Article 13. On 30 January 2021, the French Government decreed that French citizens living outside the European Union could return to France only if they had an “imperious motive”. Some French expatriates took this to the Council of State, which struck down the decree because it was in violation of Article 13 of the UDHR [15]. Secondly, as the fight against COVID-19 continues, governments that were, at the beginning of the crisis, able to take stringent measures aimed only at curtailing the spread of COVID-19, now have to take into account many different aspects, including in particular some related to maintaining economic activity. In the modern global economy, interconnections are crucial and even with the tightest of border controls, there are still many aspects of economic activity that require transborder movement of goods and, as a consequence, of those in charge of assuring this movement. Downturns in economic activity have a demonstrable and quantifiable effect on the health of populations; see, e.g., [13, 17, 23].

Thus, when taking their decisions, governments have to weigh the direct human costs of the ongoing crisis versus the societal and indirect human costs of the measures taken to curtail it. As such, understanding the role of importations and the effect of border control measures is critical. To contribute to a better understanding of these issues, we extend a model for the importation of SARS-CoV-2 to locations seeing little or no local transmission [5], to consider the risk of importing a variant to a population in which a resident virus is already circulating. This is done through three different models of increasing complexity that are schematised in Figure 1. The first model describes the spread of two variants in an isolated population. The second model adds an *importation layer* to the first model, allowing to tackle the specific roles of imported and community generated cases in local spread. The last model introduces a metapopulation-type structure to study the relationship between an exporting and an importing jurisdiction. Each model is an extension of the one before and we present and study them in succession.

**Figure 1:**
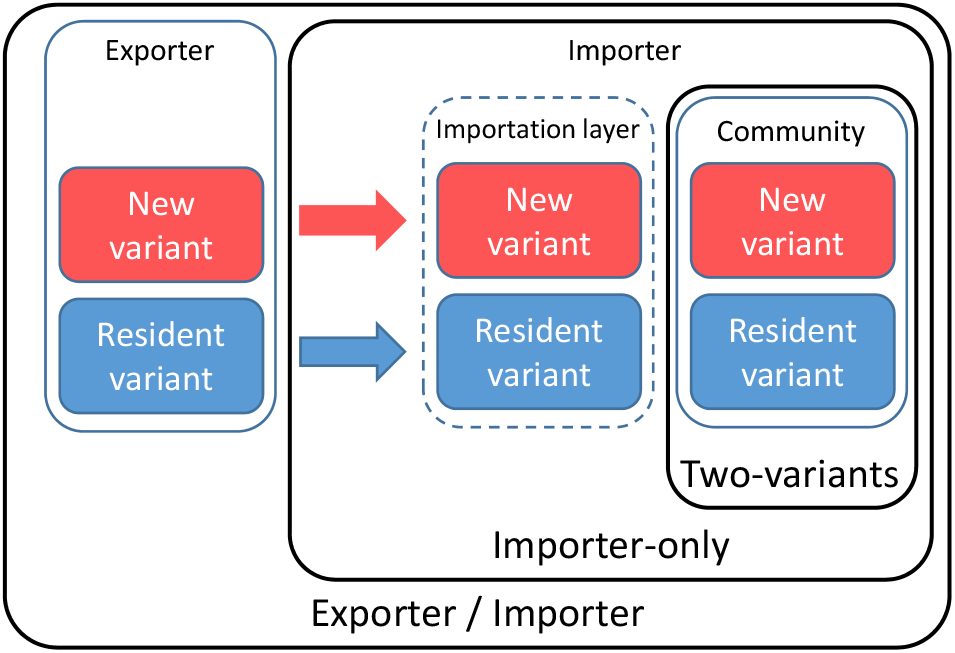
The three models under consideration : Two-variants, Importer-only and Exporter/Importer.

In this preliminary background work, we present the three models and some early mathematical and numerical investigations of their properties. Real-life work considering specific scenarii will be considered in further work. As was done in [5], while the models are formulated using ordinary differential equations (ODE), their continuous time Markov chain (CTMC) equivalents are used in practice. The advantage of using ODE is that some quantities are easy to derive in this context, such as the effective reproduction number ℛ_0_. By simulating the models as CTMC, on the other hand, we are able to track individuals as well as events and to incorporate stochasticity in trajectories.

## 2. The model for two variants

### 2.1. Case detection-based modelling

All three models in the paper are based on the compartmental model in [5], with notation adapted to better reflect the nature of the compartments. Deviating from a classic SLIAR model [6], we replace the compartments for symptomatic and asymptomatic infections, *I* and *A* respectively, by compartments for *detected* and *undetected* cases, denoted *D* and *U* respectively. This better reflects the reality of COVID-19: detected individuals are those with a positive test for the disease, whether or not they show symptoms, while undetected individuals are those who finish their infection having never been tested or having been wrongly declared negative by a test. The parameter *p* used to represent the proportion of detected cases (see Section 2.2) is thus a theoretical parameter representing testing effort given perfect knowledge of the burden of the epidemic. To precisely estimate *p*, one would indeed need, after the epidemic is over, to compare the total detected and total burdens of the epidemic, i.e., perform *a posteriori* testing of the entire population, including all deaths having occurred during the epidemic. While a precise estimation of *p* is thus nigh impossible, this parameter does provide a way to rank testing effort between different locations or groups.

Progression through undetected compartments ultimately ends in *removal*, which happens because of recovery or death, i.e., using a Kermack-McKendrick type interpretation of removal. For detected compartments, in order to potentially reconcile model outputs with data, removal is explicitly split between *recovery* and *death*; the latter is denoted *M* (for *mort* in French). As in [5], compartments *L, U* and *D* are split into sub-compartments, with two used here for simplicity, in order have Erlang-distributed times of sojourn in these types of states; see details in [9]. We call *unobserved* compartments and denote 𝒰 the latent and undetected compartments, i.e., those where individuals bear the disease but have not yet been detected or are not detected altogether.

### 2.2. Two-variants model formulation

The model for two variants in an isolated location is a straightforward extension of the compartmental model in [5], with notation modified as explained in Section 2.1. To describe the spread of two variants, the structure in the model of [5] is doubled, with one branch containing the *original* or *resident* variant, denoted with the index *O*, and another containing the *novel* or *new* variant, denoted with the index *N*. The model structure is shown in Figure 2. To keep notation consistent throughout the paper, we write these indices *O*_*C*_ and *N*_*C*_, where *C* indicates infections acquired from individuals in the community.

**Figure 2:**
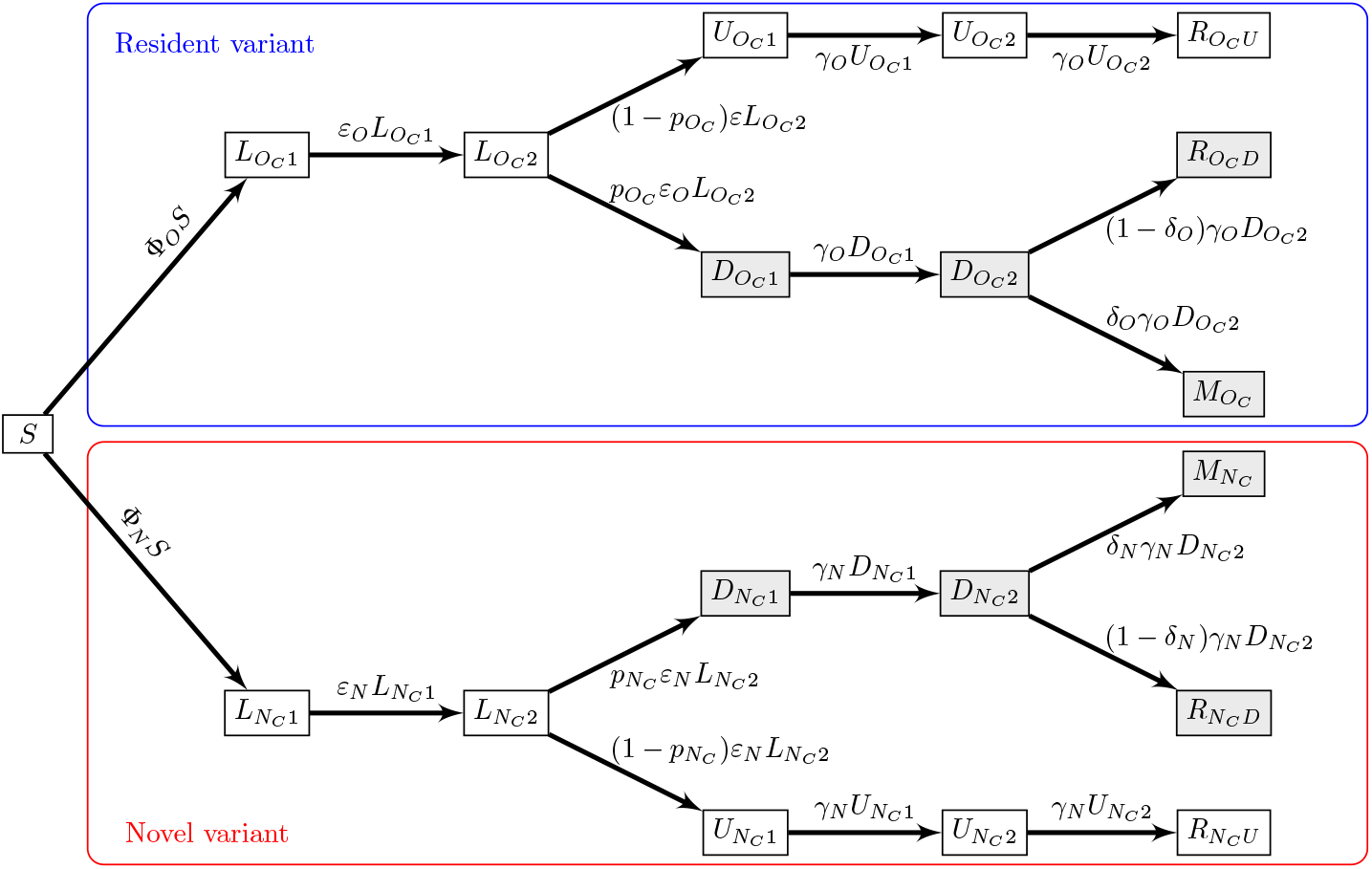
Flow diagram of the two-variants model. Shaded compartments include detected (observable or observed) cases; blue and red boxes show resident and novel variants, respectively. Φ_*O*_ and Φ_*N*_ are the forces of infection for the resident and the original variants, respectively, and are given by (A.2) in Appendix A.

The model is formulated using ordinary differential equations, which are shown in Appendix A, while simulations of the model are performed as continuous-time Markov chains. It is assumed that the period of time under consideration is sufficiently small that demographic effects do not play a role. As a consequence, the only demographic parameter used is the total population *P* in the location. Epidemiological parameters of the model are summarised in Table 1 and are allowed to vary between the original and the novel variant, to reflect that characteristics of the novel variant may differ substantially from those of the resident variant. Values indicated were chosen to reflect the general characteristics of COVID-19 rather than one of the specific situations detailed in the literature. Note that we use a rather long generation interval of 5 + 10*/*2 = 10 days compared to estimates that generally put it around 7 days [1, 18, 26, 35]. However, we are, in this preliminary work, concerned mostly with observing general trends rather than making precise predictions. The main effect of using longer generation intervals is seen when we consider quarantine later in the work, where we find quarantine efficacy lower than it would be for shorter generation intervals. Also note that most studies on disease characteristics date back to the first wave and it is not unlikely that some of these characteristics have evolved since or that they would be different for some of the variants.

**Table 1:** Parameters for variant *X* ∈ *{O, N}*.

| Parameter | Meaning | Value or range |
| --- | --- | --- |
| $\mathcal{R}_0^X$ | Reproduction number | [0.5, 3.0] |
| $\beta_X$ | Disease transmission parameter | Computed |
| $\eta_X$ | Modulation of transmissibility for latent individuals | [0, 1] |
| $\xi_X$ | Modulation of transmissibility for detected individuals | [0, 1] |
| $2/\varepsilon_X$ | Average duration of the incubation period | 5 days |
| $2/\gamma_X$ | Average duration of the infectious period | 10 days |
| $\delta_X$ | Case fatality ratio | 0.53% [29] |
| $p_{X_C}$ | Efficacy of detection in community cases | [0, 1] |

For variant *X* ∈ *{O, N}*, parameters are as follows. The rate of moving through latency (assumed to completely match incubation) is *ε*_*X*_. Because of the Erlang distribution resulting from using two compartments in *L*, 2*/ε*_*X*_ is the average duration of the latent period. Similarly, 2*/γ*_*X*_ is the average duration of the infectious period. Testing intensity is described by the parameter 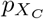 ∈ [0,1], while the case fatality ratio is *δ*_*X*_ ∈ [0, 1]. Recall that we distinguish between recovery and death only for detected individuals; for undetected individuals, both processes are aggregated within “removal” and as a consequence, *δ*_*X*_ only acts on detected individuals. Finally, *β*_*X*_ is the disease transmission parameter. In terms of units, 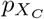 and *δ*_*X*_ are proportions, *ε*_*X*_ and *γ*_*X*_ are *per capita*, while *β*_*X*_ is *per capita per contact*.

### 2.3. Mathematical properties of the two-variants model

We first consider properties of the ODE model, since some of these properties carry through to the CTMC model. First, there are two types of reproduction numbers; see Appendix A.2 and Appendix A.4 for details of their computation. The variant-specific reproduction number is given, for variant *X* ∈ *{O, N}*, by

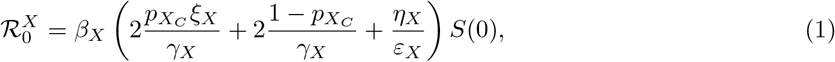

where *η*_*X*_ and *ξ*_*X*_ are the modulation (typically attenuation) factors for transmission by incubating and detected cases and *S*(0) is the susceptible population at the initial time, computed by subtracting the total non-susceptible population from the total population *P*. There is also a “system-wide” reproduction number,

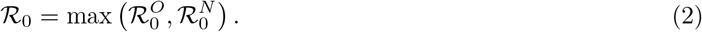

When considering a single variant in isolation, it is possible to derive a final size relation, i.e., to obtain the overall attack rate of the variant should the other variant not be present; see Appendix A.3.

### 2.4. Setting up simulations of the two-variants model

The free parameters 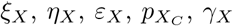 and 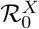 are estimated from the literature or by calibrating model responses to data. As indicated earlier, our aim in this work is not to be extremely precise with regards to parameter values but rather to use values compatible with the general characteristics of COVID-19. The disease transmission parameter *β*_*X*_ for variant *X* ∈ *{O, N}* is obtained from the reproduction number 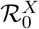 for this variant by solving (1) for *β*_*X*_ as a function of 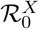, finding the transmission coefficient for variant *X* as

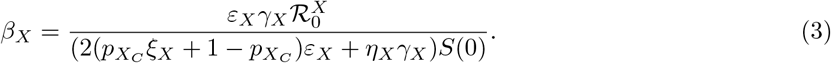

For simplicity, in this introductory work, we take the values of the rate of progression through the incubation and infectious periods and the disease induced death rate to be constants and vary only the reproduction number 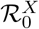 (and through it *β*_*X*_) and the efficacy of detection 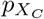.

We use the `R` package `GillespieSSA2` to perform numerical simulations of the CTMC. The package allows to track individual firings of the CTMC, i.e., the reactions that activate to change the state of the system. We utilise this feature and, in the remainder of the text, denote *Y* →*Z* the event in the CTMC that results in the switch of an individual from state *Y* to state *Z*. If transitions are “obvious”, some information is omitted; for instance, the transition 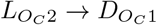 is written 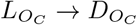, since it is evident from model structure that transition from latent to detected occurs between the second latent compartment and the first detected compartment (Figure 2). We denote #(·) the number of events of a certain type per unit time; e.g., # 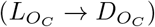 is the number per unit time of cases bearing the resident variant detected in the community. When considering different types of events, we use the *logical or* notation ∨; e.g.,

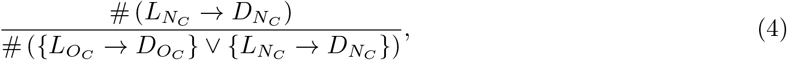

is the fraction of newly detected cases that are bearing the novel variant. Finally, new infections are denoted in a manner making explicit what type of individuals are involved. For instance, 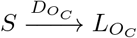 denotes new infections in the community by detected individuals bearing the resident variant, while 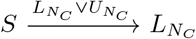 are infections by unobserved (latent or undetected) individuals.

To ensure that simulations run in a way where the start of importations of the new variant has a direct effect on the system, we first run the system without the new variant over a burn-in period of three weeks. The initial condition for the original variant used for the system with the new variant is the state of the system after this initial burn-in period. The original variant is assumed to propagate with 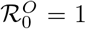, while the reproduction number for the novel variant used is 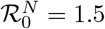 unless otherwise indicated.

In the figures throughout the remainder of the text, when a range is shown, it is the range containing *x*% of simulation results at each time point (typically 90% or 95%), between the (100 −*x*)*/*2-percentile and 100 − (100 −*x*)*/*2-percentile of values for that time point. Because each realisation of the CTMC has a different sequence of jump times, envelopes as well as mean and percentiles are obtained by interpolating each solution on a fine time grid.

### 2.5. Numerical investigation of the two-variant model

Figure 3 shows the 95% range and the mean of 1,000 simulations where *P* = 100, 000, 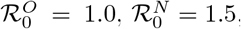, *ξ*_*O*_ = 0.15, *ξ*_*N*_ = 0.3 and there is initially a single individual in the 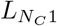 compartment. The top panel shows the numbers of infections that are with the resident variant (blue) and the novel variant (red). The slow decrease of the average number of new infections with the resident variant is due to the fact that *S*(*t*) decreases with time. This implies that the effective reproduction number 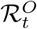 for resident variant *O*, obtained at time *t* by replacing *S*(0) with *S*(*t*) in (1), decreases. As 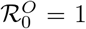 initially, this means 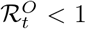 for all *t >* 0.

**Figure 3:**
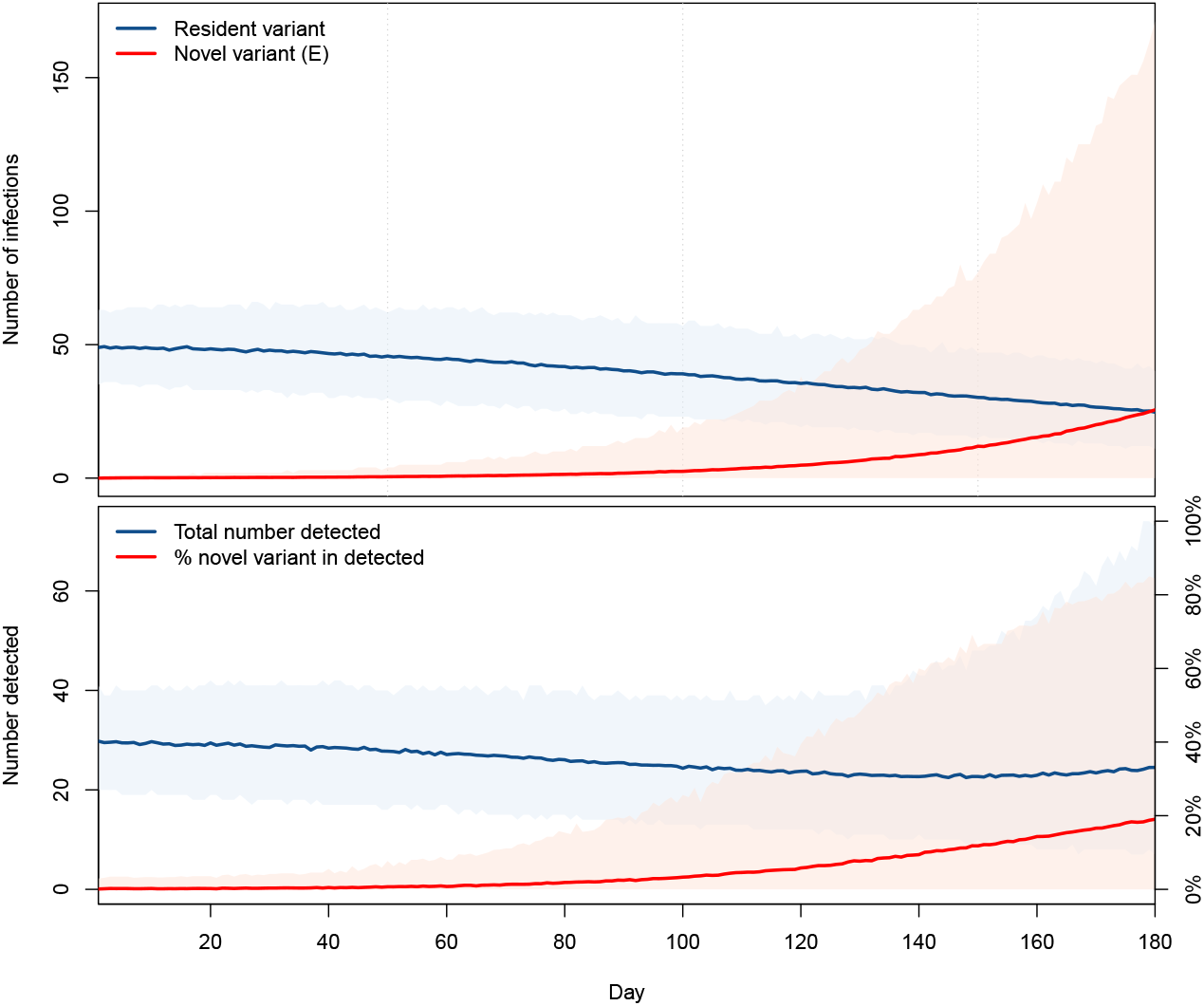
Top: number of infections with the resident (blue) and novel (red) variants in an isolated location. Bottom: total number of new cases detected (blue, scale on left axis) and percentage of these cases that are with the novel variant (red, scale on right axis). Ranges are 95% of the solutions, lines represent the mean of all solutions.

The bottom panel in Figure 3 shows the corresponding total number of detected cases, i.e., the denominator in (4), and the percentage of novel variants in these detected cases given by (4), when the percentages of resident and novel variants detected are, respectively, 60% and 40%. By the design of the model, the time to detection is underestimated, i.e., we find detection times earlier on average than they should be. Indeed, recall that a case is detected, for variant *X* ∈*{O, N}*, when a transition *L*_*X*_ → *D*_*X*_ occurs, whereas in “real life”, this transition can happen at any point during the transition through compartments downstream from latent compartments in the flow diagram of Figure 2. Thus, detection timing results presented in this work are best-case scenarios where detection occurs at the end of the incubation period.

The initial number of individuals infected with the new variant greatly influences the latter’s capacity to become established in the population. We illustrate this in Figure 4, which shows the dependence of the percentage of simulations seeing an extinction of the novel variant in the six months time window considered on the number of individuals initially infected with that variant. Parameters are as used in Figure 3. For consistency of results, we assume that at time *t* = 0, all individuals infected with the novel variant are in compartment 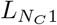. Also shown on this figure is the probability 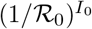 of a minor epidemic given by Whittle’s formula [34], where *I*_0_ is the number of individuals initially infected with the disease. Whittle establishes that when ℛ _0_ *>* 1, there can only be minor or major epidemics, where minor epidemics correspond to those where we observe significantly fewer cases and disease extinction generally occurs more quickly than in the case of a major epidemic [11, 31]. In the present case, since 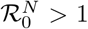, the complement to the probability of a minor epidemic is the probability of a major epidemic [24, 31, 34], corresponding here to the case where the novel variant does not become extinct. In [5], the latter probability was used to characterise *critical successful importations*.

**Figure 4:**
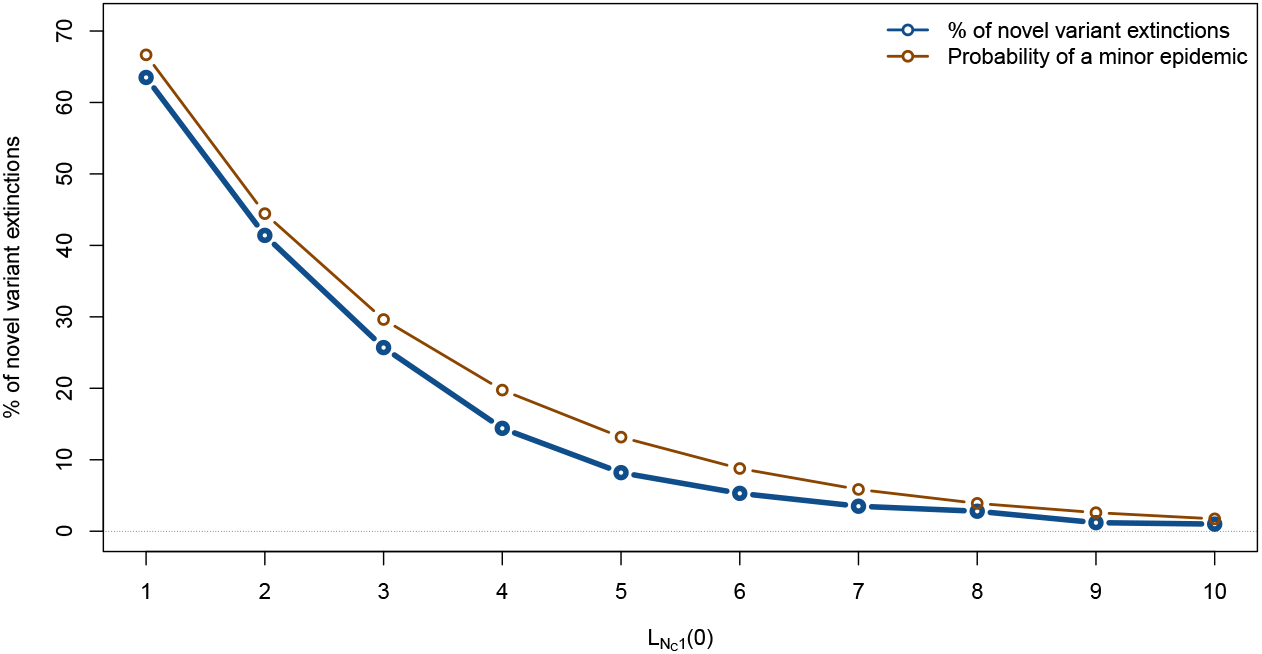
Percentage of 1,000 simulations seeing extinction of the novel variant within a six months period as a function of the initial number of individuals in 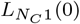, with all other compartments for the novel variant starting empty. Also shown is the theoretical value of the probability of a minor epidemic obtained by using Whittle’s formula [34], which here roughly approximates the probability of novel variant extinction; see text for details.

## 3. The importer-only model

To study the role of importations and the effect of control measures implemented to limit them, we now add compartments that contain all individuals bearing the disease and arriving in the location of interest from outside the location of interest. We focus here on the dynamics of importation within the location of interest and thus call this the *importer-only* model.

### 3.1. Formulation of the importer-only model

We consider a model with the same structure as in Section 2 but subjected to importations. We adopt the same language as in [5]: *importations* are all events when someone infected with the disease in another location arrives in the location of interest; *successful importations* are those importations that result in the transmission of the disease to at least one individual in the location of interest, i.e., success is from the perspective of the pathogen.

We assume that importations can only be of individuals incubating with the disease (*L* compartments) or with an undetected active infection (*U* compartments); individuals with detected active infections are not able or allowed to travel.

In order to distinguish between infections happening because of contacts with imported and community cases, we introduce an *importation layer*. Individuals in the importation layer and in the community are distinguished using indices *I* and *C*, respectively. Thus, for variant *X* ∈ *{O, N}*, cases are distinguished by the indices *X*_*I*_ and *X*_*C*_, depending on whether infection was acquired by contact with someone infected in another location or in the community. Each disease state is therefore indexed by of one of four types: resident variant acquired in the community *O*_*C*_, imported resident variant *O*_*I*_, novel variant acquired in the community *N*_*C*_ and imported novel variant *N*_*I*_.

By model construction, new infections in the importation layer can only originate from an importation, i.e., there is no within-group transmission of the pathogen there. These importations occur at Poisson distributed times with rate constant *λ*_*X*_ for variant *X* ∈ *{O, N}* and enter compartments 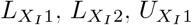 and 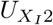 with probabilities 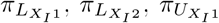 and 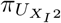, respectively, where 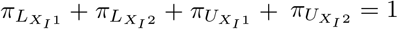. Individuals in the importation layer make contacts with individuals in the community freely.

We assume that detection efficacy can differ between imported and community cases for a given variant. Indeed, individuals entering a jurisdiction are often subject to different detection protocols than individuals already in the jurisdiction. For instance, Canada and the United Kingdom have been using “day 0” testing, where an individual needs, additionally to a negative PCR test less than 72 hours before undertaking travel, to submit to a PCR test upon entry into the jurisdiction.

As is the two-variants model in Section 2, we assume that the total population in the location of interest is *P*. The model structure is shown in Figure 5.

**Figure 5:**
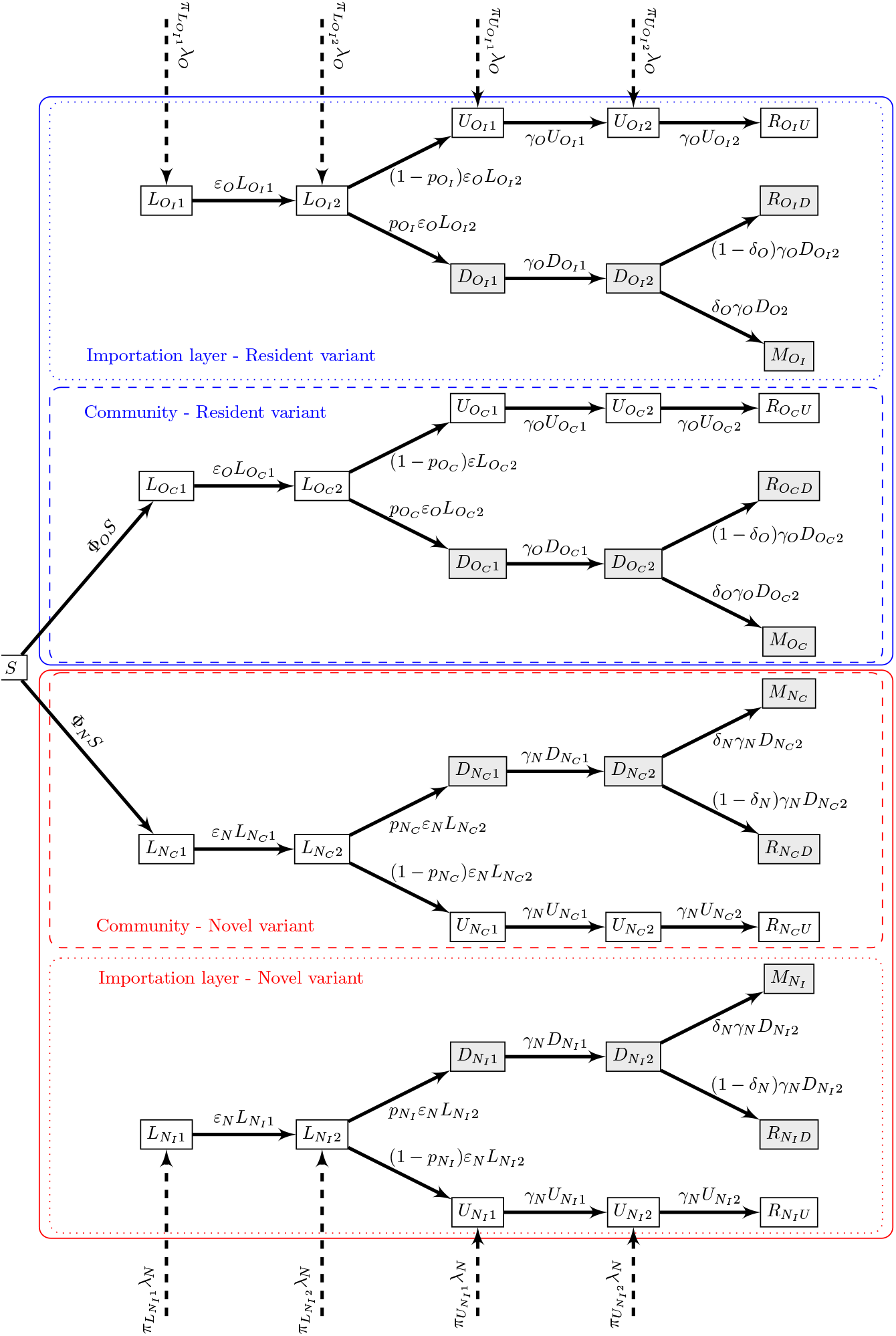
Flow diagram of the importer-only model. Shaded compartments are detected; blue and red boxes show resident and novel variants, respectively. For variant *X* ∈ *{O, N}*, Φ_*X*_ is the force of infection (B.2) in Appendix B.

### 3.2. Properties of the importer-only model

Because importations always enter and remain in the importation layer, *P* is constant for the community but the total population in the location of interest is increasing.

The importer-only model does not admit a disease-free equilibrium, because of the inflow of individuals into infected compartments. As a consequence, it is not possible to compute a reproduction number for the entire system. See, e.g., [2] for a discussion of this issue and some leads into addressing it. It is still possible to use, and we do, the formula (1) for the reproduction number for a single variant without importation, but this reproduction number must be understood to apply in this specific situation.

### 3.3. No travel control

Let us start with the case with no travel control, to understand the baseline response of the model. To visualise the role of importations, we compare solutions of the model without importations of Section 2 with solutions of the importer-only model, for increasing values of the rate of importation *λ*_*X*_. In Figure 6, we focus on incidence of the novel variant, when 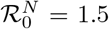. As expected, importations speed up spread: the base case without importations (dashed line) sees the slowest rate of increase of the incidence of the novel variant, while increasing importation rates result in increasingly fast epidemics. Simulations shown are run with an importer population of 100,000 and at high importation rates, we observe that the epidemic peak is reached within the six-months period of interest, much earlier than for lower or zero importation rates.

**Figure 6:**
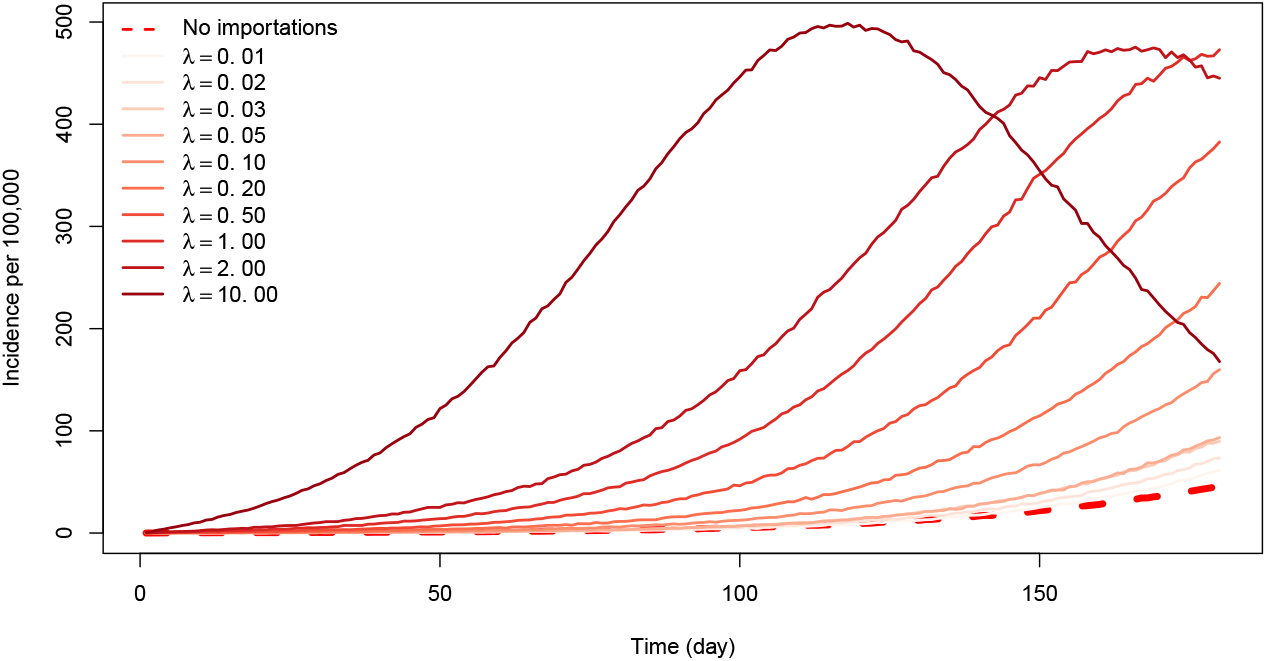
Incidence of the novel variant in the two-variants model without importation (dashed line) and in the importer-only model with increasing importation rates *λ* := *λ*_*O*_ = *λ*_*N*_. Here, 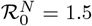.

To investigate the likelihood of detection of new variants, let us consider the pendant of the lower panel in Figure 3, i.e., the evolution of the fraction of detection of the novel variant among all detections,

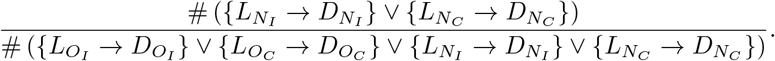

The evolution of this quantity is shown in Figure 7, when the percentages of resident and novel variants detected are, respectively, 60% and 40%. For the parameters used, we observe that the introduction of the novel variant does not have an immediate effect on the total number of detected cases (blue curve); it takes almost 100 days until this curve starts to increase. On the other hand, detection of the novel variant occurs more rapidly here than it does in the isolated case with no importation of the lower panel of Figure 3. Note, however, that while the mean percentage of novel variant among all detected cases rises faster, there remain quite a sizeable fraction of realisations for which this percentage is low until about day 60. As a matter of fact, until around day 25, the median value of the percentage (shown by the dashed black line tracking close to the mean percentage in Figure 7) is 0, meaning that half of the realisations see no detection of the novel variant at all until this point.

**Figure 7:**
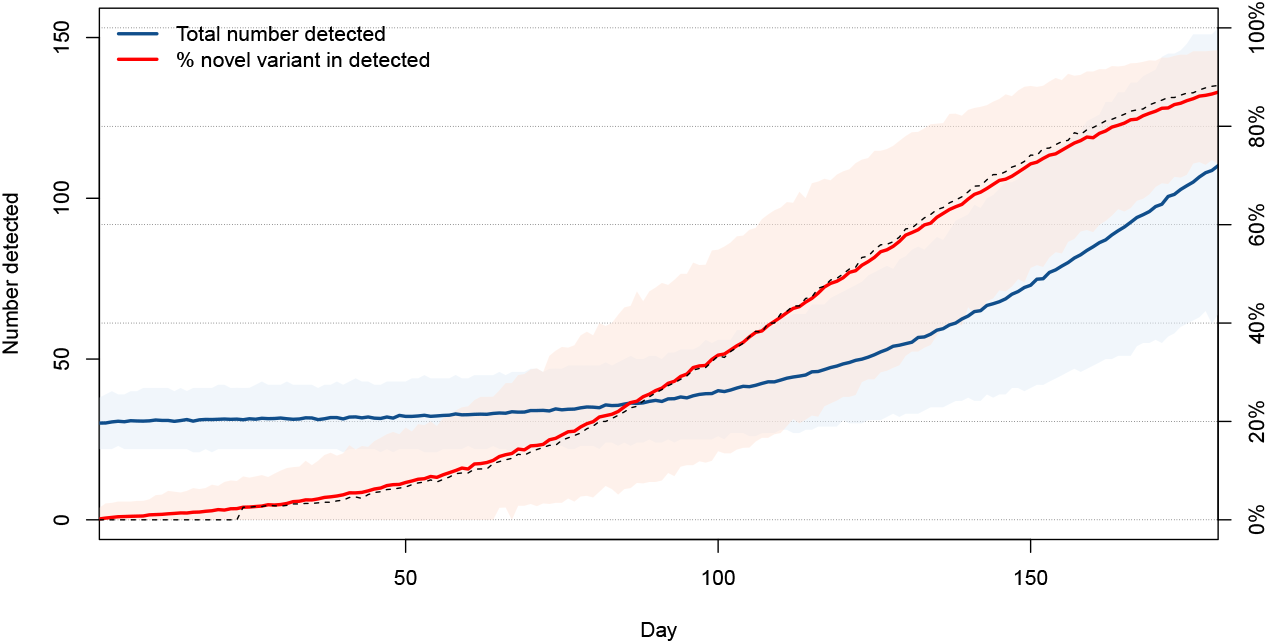
Mean total number of new cases detected (blue, scale on left axis) and mean percentage of new variants detected within these (red, scale on right axis). Also shown in black is the median number of detections of the novel variant. Here, *λ*_*O*_ = *λ*_*N*_ = 1*/*2.

### 3.4. Effect of quarantine

To consider the effect of quarantine, we use the method developed in [5] and detailed in Appendix B.2. We proceed in several steps illustrated in Figure 8. We assume that what is happening in the rest of the world translates, for variant *X* ∈ *{O, N}*, into an importation rate *λ*_*X*_ with distribution of states *C*_*X*_ (0), where 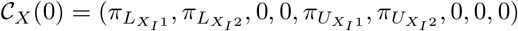.

**Figure 8:**
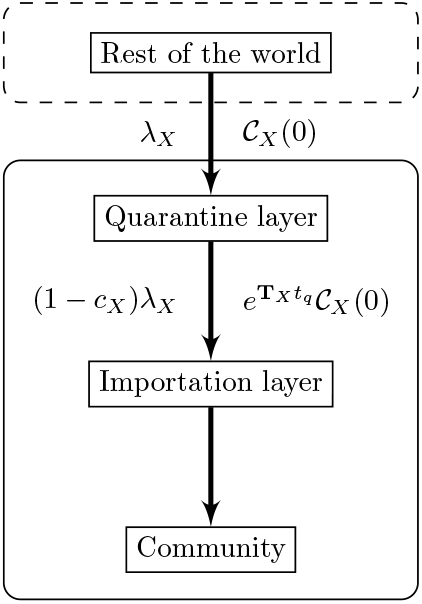
Setup of the quarantine simulations.

Here, for simplicity, we assume that not-yet-detected or undetected cases are equally likely, i.e., 𝒞 _*X*_ (0) = (1*/*4, 1*/*4, 0, 0, 1*/*4, 1*/*4, 0, 0, 0). From this and model parameters, we deduce both the quarantine efficacy *c*_*X*_ and the distribution of probability of being in the different compartments given by (B.4). This is the output of the quarantine layer and is fed into the importation layer. Using the indicator vector *u* defined in Appendix B.2, inputs to 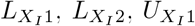 and 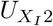 in the importation layer occur at rate given by the nonzero entries in 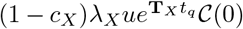, where 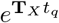 are the probabilities of transition between states for individuals over *t*_*q*_ units of time and *t*_*q*_ is the quarantine duration.

In Figure 6, we observe that importations can be disastrous in terms of overall disease burden. In order to evaluate the impact of quarantine, we compare the outcome of simulations without quarantine as in Figure 6 and with three durations of quarantine that have been used by different countries, *t*_*q*_ = 7, 10 and 14 days. One thousand simulations are run for each combination of importation rate and quarantine duration. For each simulation, we compute the numeric final size of the novel variant by counting how many infections with the novel variant take place over the period of interest of six months. Figure 9 then shows the median value of the final sizes.

**Figure 9:**
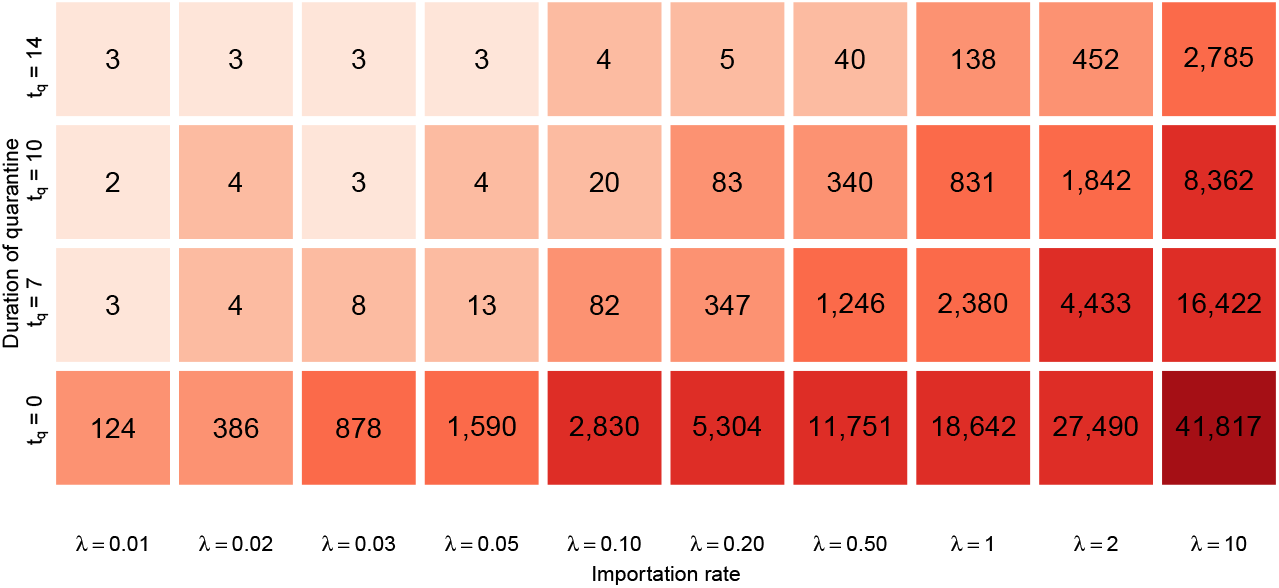
Median over 1,000 simulations of the final size of the novel variant over the period of interest of six months, as a function of the importation rate *λ* := *λ*_*O*_ = *λ*_*N*_ and the duration *t*_*q*_ of quarantine (in days). The bottom row corresponds to final sizes of the solutions shown in Figure 6.

### 3.5. Effect of complete border closures

To evaluate the role of complete interruption of travel, we proceed to the following experiment. We choose a day *t*_*d*_ for the start of travel control. On the interval [0, *t*_*d*_], we run the system as in Section 3.3. Then, starting on day *t*_*d*_, full travel interruption is implemented by setting *λ*_*O*_ = *λ*_*N*_ = 0.

Figure 10 shows the mean trajectories of 1,000 simulations, i.e., the mean numbers of transmission events, in the same setting as Figure 7 and with a moderate importation rate *λ*_*O*_ = *λ*_*N*_ = 1*/*2 prior to border closures, for values of *t*_*d*_ = 10, 20, …, 180. (The case *t*_*d*_ = 180 means there is never an interruption of travel.)

**Figure 10:**
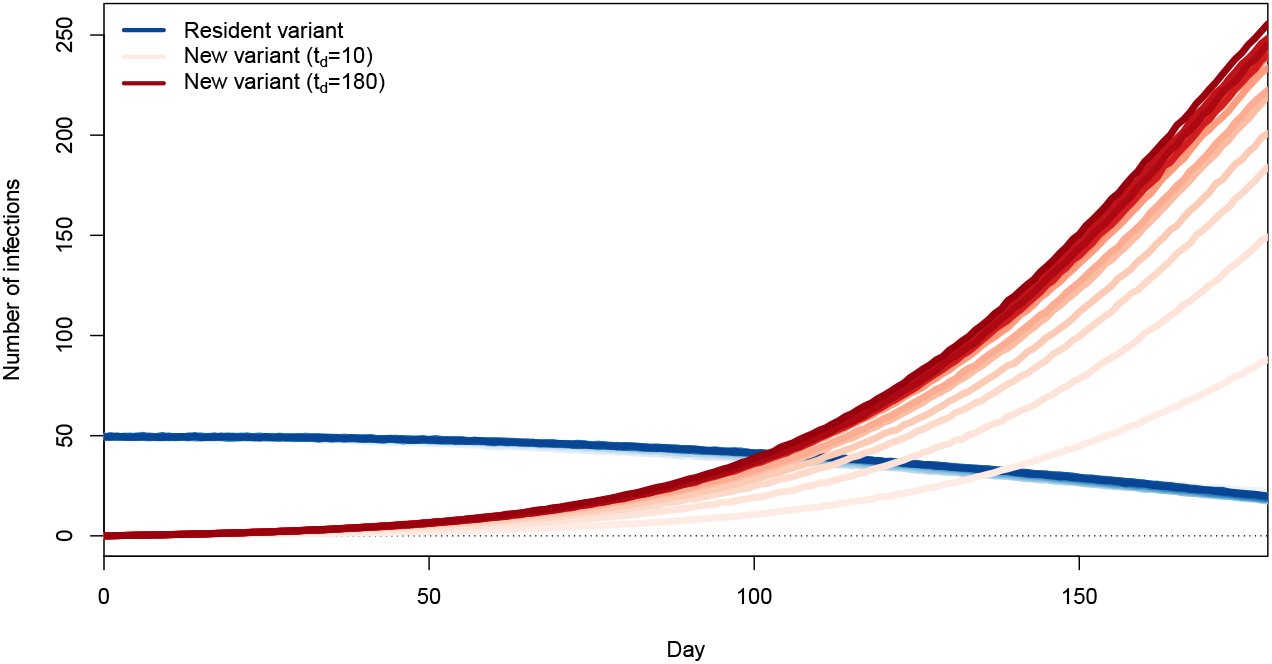
Number of transmission events involving the original (blue) and novel (red) variants, with a complete interruption of importations occurring at *t*_*d*_ = 10, 20, …, 180 days. Scales (most not shown) range from lightest when *t*_*d*_ = 10 days to darkest when *t*_*d*_ = 180 days. Each curve is the mean of 1,000 simulations.

We see that trajectories more advantageous in terms of control of the variant are those corresponding to *t*_*d*_ = 10 and 20 days, with other trajectories being roughly similar to one another. Interruption of importations does not affect the already spreading resident variant. Thus, it appears that unless they are implemented very soon after the novel variant starts being imported, travel restrictions play very little role. To understand why this is, consider the main panel in Figure 11. There, we observe that community transmission quickly becomes the main source of transmission of the new variant, while transmission from imported cases to the community remains very low. Thus, very little seeding by imported variant cases is required to start a local outbreak. Early efforts (*t*_*d*_ = 10 or 20) substantially delay passage times through set incidence values. The likely reason for this is that for such values of *t*_*d*_, we fail to import a case. Indeed, from the model, the probability of no successful variant introduction is approximately exp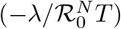 over the period [0, *T*]. This value becomes less than 5% when 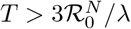 The mean time to introduction of the first novel variant is 1*/λ* and mean time to the first successful introduction is 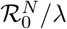. Longer times to travel shutdown, starting with *t*_*d*_ = 30 days, seem to change very little to the outcome.

**Figure 11:**
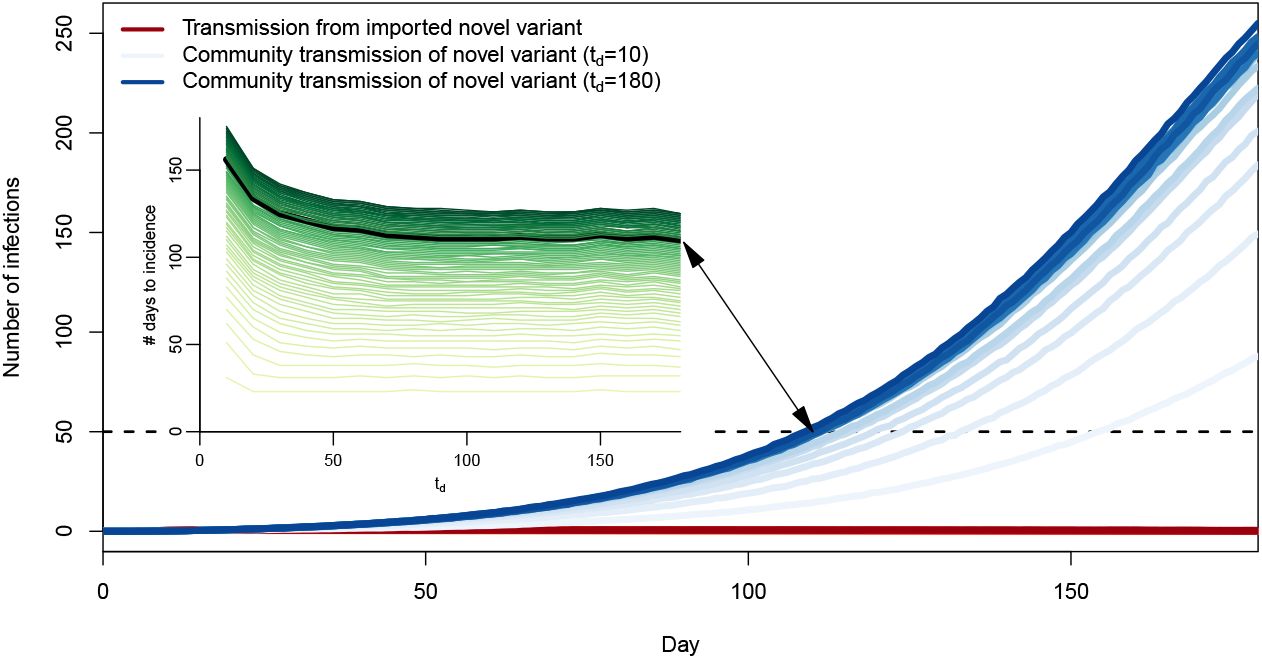
Number of transmission events involving importation cases bearing the new variant 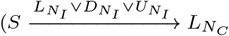, red) and transmission of the new variant within the community 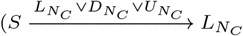, blue), when a complete interruption of importations occurs at *t*_*d*_ = 10, 20, …, 180 days. Scales (most not shown) range from lightest when *t*_*d*_ = 10 days to darkest when *t*_*d*_ = 180 days. Each curve is the mean of 1,000 simulations. Inset: mean first passage time (MFPT) to a prescribed incidence value as a function of the number *t*_*d*_ of days from the start of importations to complete travel interruption. Lightest coloured curve: MFPT to an incidence of 1; darkest colour: MFPT to an incidence of 87.

To confirm this, consider the inset in Figure 11, which shows the day at which each of the blue curves in the main panel in the figure passes through a given incidence value, i.e, the mean first passage time (MFPT) through that value. Curves are ordered and coloured from the MFPT for an incidence of 1 at the bottom in light green to the MFPT for an incidence of 87 at the top in dark green, which is the maximum incidence at 180 days along the blue curve for *t*_*d*_ = 10. The example of the MFPT through an incidence of 50 is shown, with the point where the curve for community transmissions with *t*_*d*_ = 180 crosses an incidence of 50 highlighted together with the corresponding point on the MFPT at incidence 50.

One way to read the MFPT curves is as follows. Set a target maximum daily incidence and pick the green MFPT curve corresponding to this value. Its right endpoint is the situation where no action is ever taken, i.e., *t*_*d*_ = 180. Following the curve left, we observe that there is initially virtually no change, meaning that reacting with this delay achieves almost no benefit in timing. Then, as *t*_*d*_ decreases more, the curve starts to increase, meaning that time was “bought”. Observe that even for high thresholds, the change is minimal unless one is able to react within two months of the first potential importation. Further, as the threshold is decreased, so is the value of *t*_*d*_ for which meaningful time delays are achieved, to the point that for the lowest thresholds, only *t*_*d*_ = 10 achieves more than a two day increase in MFPT.

Contrary to the base case of Section 3.3 and to the case with quarantine of Section 3.4, it is possible here for the novel variant to become extinct during the period of interest, since there are no new importations after *t*_*d*_. For the simulations shown in Figure 11, such extinctions occur 209, 46 and 11 times (out of 1,000 simulations each) for *t*_*d*_ equal to 10, 20 and 30 days, respectively, and never for *t*_*d*_ *≥*40. This further confirms that the (time) window of opportunity during which complete border closures can be expected to contribute substantially to the curtailing of the disease is quite narrow.

This is all the more relevant when one considers the process for declaring an emergent novel variant a VOI or VOC, as it is indeed unreasonable to think that a remote public health authority would act to block travel from a country before a variant is declared a VOI/VOC by local authorities there. There is no consensus on the definition of VOI or VOC; the following is the one proposed by WHO in February 2021; see also the already cited US CDC definition [32]. A SARS-CoV-2 isolate is a variant of interest (VOI) if it is phenotypically changed compared to a reference isolate or has a genome with mutations that lead to amino acid changes associated with established or suspected phenotypic implications AND has been identified to cause community transmission/multiple COVID-19 cases/clusters, or has been detected in multiple countries; OR is otherwise assessed to be a VOI by WHO in consultation with the WHO SARS-CoV-2 Virus Evolution Working Group. In view of this definition, the time it takes before a variant is declared *of interest* will vary based on local authorities as well as the efficiency of local surveillance. This suggests that there is a lower bound for *t*_*d*_ that may well be greater than or equal to the threshold of 30 to 40 days after which the novel variant is shown to almost always establish.

### 3.6. Comparing the effect of quarantine and border closures

To compare the effect of quarantine and complete travel interruptions, we choose to consider the final size of infections with the novel variant using both modalities of importation control. The result is shown in Figure 12. Parameters used are as in Sections 3.4 and 3.5 and we consider the moderate value of *λ*_*O*_ = *λ*_*N*_ = 1*/*2, i.e., one importation event on average every two days.

**Figure 12:**
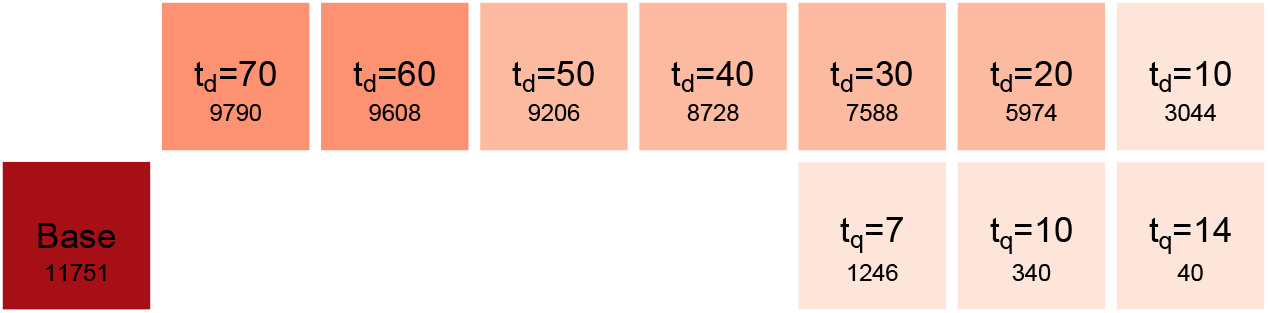
Final sizes of infections with the novel variant over a six months period, in the base case (importation without control, left lower row), with quarantine (right lower row) and with complete travel interruptions (upper row). The number indicated below the type of control is the final size.

To understand why even a ten days delayed travel interruption is less effective than a 7 days quarantine, recall that the numbers presented are averages over 1,000 simulations. We saw in Section 3.5 that with complete travel interruptions after 10 days, about 20% of simulations saw complete extinction of the novel variant over the 180 days period of interest. However, other than these favourable simulations, other simulations with *t*_*d*_ = 10 had on average 5 or 6 introductions before complete shutdown of importations. On the other hand, with the model parameters used, a quarantine with *t*_*q*_ = 7 is 55% efficacious, implying a multiplication of the mean time between importations by 1*/*0.45 *≃* 2.22 and that instead of an importation rate *λ*_*X*_ = 1*/*2, the location is subject to importation rates of 0.225.

## 4. The exporter-importer model

In our considerations in Section 3, importation rates *λ*_*O*_ and *λ*_*N*_ are held constant throughout simulations. In Section 3.4, they are modulated by quarantine, while in Section 3.5 they are set to zero after *t*_*d*_ days to mimic border closures. However, they do not depend on the evolution of the epidemic situation in locations from which imported cases originate.

As a consequence, when considering the number of infections involving imported infectious individuals versus those in the community, we find that the number of new infections generated by imported cases is essentially a function of the importation rates *λ*_*O*_ and *λ*_*N*_ and remains roughly constant through time.

To better understand the role of detection in relation to border closures, we therefore extend the model to take into account the epidemic situation in the location where imported cases originate from.

### 4.1. Formulation of the exporter-importer model

We extend our study to the mechanism generating the importations received by the model in Section 3. For this, we derive a metapopulation-type model [4] with only two spatial locations: an *exporter*, where amplification of the novel variant is taking place, and an *importer*, which uses the model formulated in Section 3.

We use the superscript *E* to indicate state variables and parameters in the exporting jurisdiction. In the exporter, we use a replica of the model in Figure 2 for propagation of two variants within a community in Section 2. We suppose that individuals in compartments 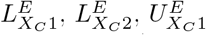 and 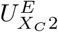, *X* ∈ *{O, N}*, in the exporter travel to the importer at the per capita mobility rates *λ*_*X*_. As previously, we assume detected individuals do not move between locations. Most parameters are assumed to take the same values as in the importer, except for the contact parameter *β* and the proportion of detection *p*. The total population of the exporter is *P*^*E*^ initially, while the total population of the community in the importer is fixed and equals *P*. See Figure 13, which details the model structure for one of the variants *X* ∈*{O, N}*. Model equations for the ODE exporter-importer model are given in Appendix C.1.

**Figure 13:**
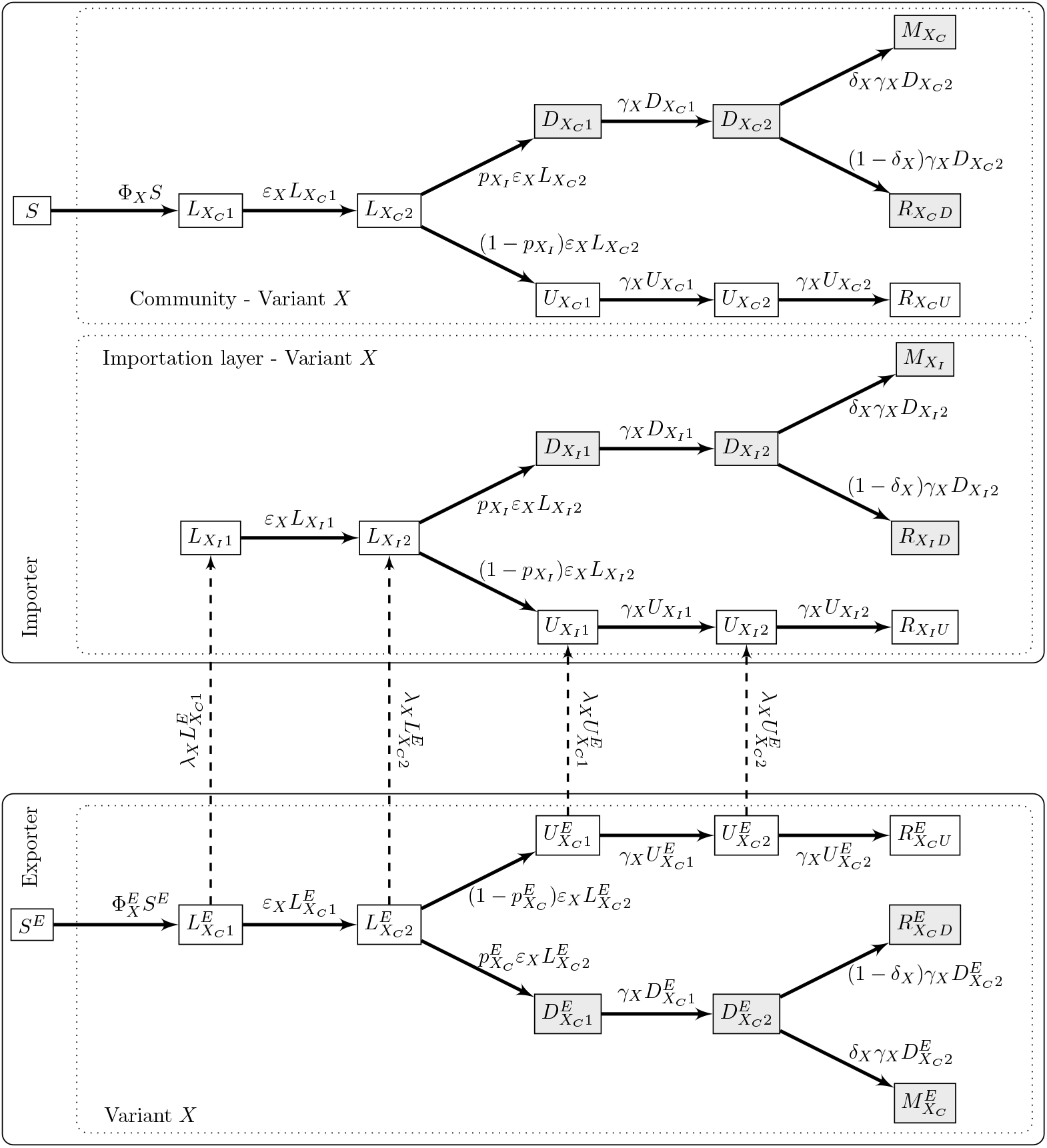
Export-import model for one variant *X* ∈ *{O, N}*. Recall that *S* and *S*^*E*^ are also connected to the other variant *Y* ∈ *{O, N} ≠ X*.

### 4.2. Properties of the exporter-importer model

Taken together with individuals in the importation layer of the importer, the total number *P*^*E*^ of individuals in the exporter is constant, although the population in each patch varies. (Note that this includes death.) The total population in the community in the importer is also constant.

One major difference between the import-only and export-import models is that because importations are not considered in the exporter, the export-import model behaves like a regular metapopulation model and has, in particular, a disease-free equilibrium. This implies that it is possible to compute a basic reproduction number for each variant and for the whole system.

However, one can also consider the exporter in isolation from the importer, since the latter plays no role in the dynamics of the former. This is useful to set the reproduction number in the exporter patch. The computation in Appendix A.2 is virtually unchanged in this case, with the only difference being that matrix **V**_*X*_ takes the form

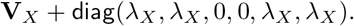

This changes the expression for 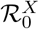 as follows; for variant *X* ∈ *{O, N}* in the exporter patch,

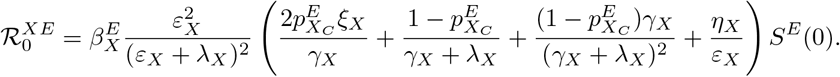

In simulations, the value of 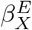 is set by solving this formula for a given 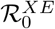, giving a complicated expression that is not shown here.

### 4.3. Sample behaviour of the exporter-importer system

The set up of numerical investigations of the exporter-importer system is similar to earlier models. To compute the movement rate *λ* from exporter to importer, we proceed as in [8]: we consider the evolution of the number *P*^*E*^ of individuals in the exporter due to movement of passengers from the exporter to the importer. Over a very short time period, we can assume that there are no other significant sources of change to *P*^*E*^, so that (*P*^*E*^) *′* = *−λP*^*E*^. Solving this simple linear ODE for one day and equating *P*^*E*^(0) *− P*^*E*^(1) = Δ_*E*_, where Δ_*E*_ is the number of passengers travelling from the exporter to the importer in one day, we find *λ* = *−* ln(1 *−* Δ_*E*_*/P*^*E*^).

Note one fundamental difference between the importation rates used in the second model (Section 3) and those used here. In the second model, the effect of importation rates is *additive*, in the sense that because of competing risks, the importer population is subject to importations at the combined rate *λ*_*O*_ + *λ*_*N*_. Here, on the other hand, the rate *λ* derived above applies *per capita* to the entire population *P*^*E*^ in the exporter patch. Therefore, in the present model, *λ*_*O*_ and *λ*_*N*_ are modulations of this rate. If they are taken equal to *λ* (which is what is typically done), travel takes place at the same rate for all individuals regardless of their epidemic status. Values of *λ*_*O*_ and *λ*_*N*_ smaller than the general *λ* may be used for instance to indicate that negative PCR tests are required for travel.

In Figure 14, we show results when the two locations have the same population of 1 million people and for *per capita* movement rates from the exporter to the importer corresponding to 500 and 5,000 passengers per day. A 21-days burn-in period is used prior to the dates shown, in which it is assumed that the resident variant is propagating in both the exporter and the importer with 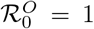, while the novel variant is propagating only in the exporter with 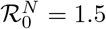, starting the burn-in period there with a prevalence equal to that of the resident variant.

**Figure 14:**
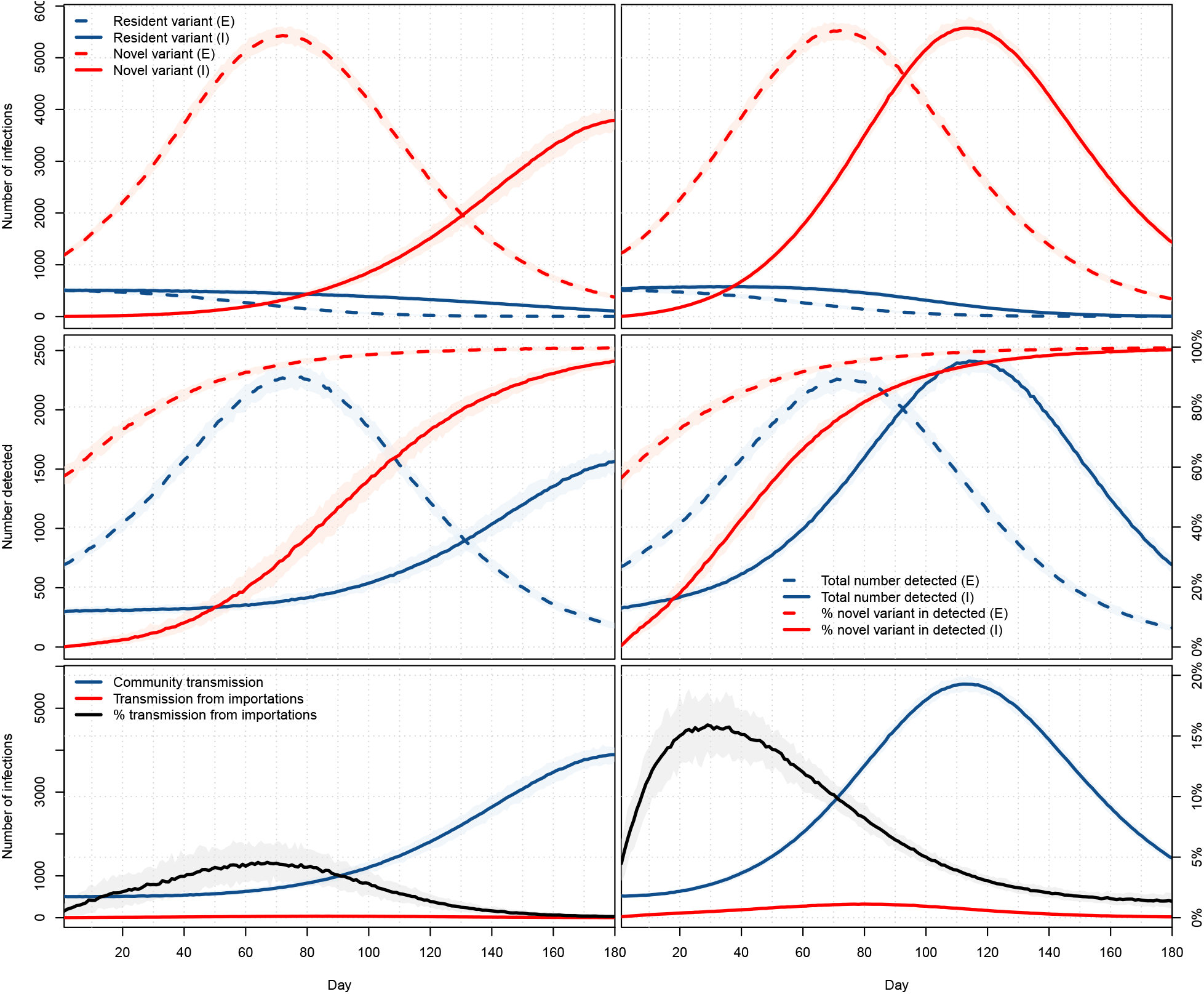
Top: number of infections with the resident (blue) and novel (red) variants in the exporter (dashed) and importer (plain). Middle: total number of detected cases (blue) and percentage of these detected cases that are with the novel variant (red) in the exporter (dashed) and importer (plain). Bottom: Number of infections in the importer patch resulting from contacts with individuals infected in the community (blue) or in the exporter patch (red). Both locations have a population of 1 million. Left column: travel of a maximum of 500 passengers per day; right column: travel of a maximum of 5,000 passengers per day. All panels: means of 1,000 simulations and ranges of 95% of these simulations.

The inflow numbers used represent 0.05% of the population per day for the low inflow case and 0.5% of the population per day for the high inflow case. To get a sense of the validity of such numbers, let us compare this with international entries into Canada, for which data is available [30]. In 2019, there were a total number of 96,810,028 people-entries into Canada, i.e., roughly 0.7% of its population per day. From April 2020 to February 2021 inclusive (the latest data available at the time of writing), the number of international people-entries dropped to 10,105,865, roughly 0.08% of the population per day. Thus the situation shown in Figure 14 is roughly consistent with unabated (right column) and abated (left column) movement rates.

The first row in Figure 14 shows the dynamics of infection of the two variants in the two locations. At low *per capita* importation rates, the peak of infections, due entirely to the novel variant, is delayed by about 100 days in the importer. This delay falls to about 40 days for regular *per capita* importation rates.

This delay is also observed when considering detections, which are shown in the second row of Figure 14. Note one interesting difference in these compared to Figure 7: even in the case of low importation rates, the percentage of detections of the new variant rises more quickly than it did there. In practice, though, the build-up of transmissions of the novel variant is more progressive. Indeed, recall that in Figure 14, we have assumed that the variant was completely absent from the importer while it was already spreading at high levels in the exporter, to the point that it already accounts for about 60% of the detected cases in the exporter at time *t* = 0.

Consequently, the influence of imported cases on the dynamics is also more important, as can be seen from the third row in Figure 14. In the case of pandemic-era mobility rates, the percentage of transmissions in the importer resulting from contacts with someone infected in the exporter (i.e., an individual in the importation layer of the importer) reaches a peak mean of about 4.5% at 67 days. When mobility is similar to what it was in the pre-pandemic era, this peak reaches 15.9% much sooner, after 29 days. Interestingly, in both instances, this peak precedes the peak in novel variant infections in the exporter, with the precedence increasing together with the mobility rate.

## 5. Discussion

We present three increasingly complex models for the importation and spread of two variants of COVID-19, which we specialise to the context of spread of a novel variant in a population already experiencing spread of another variant. These models are, at present, not tailored explicitly to data, but in view of existing data on the spread of variants, we are satisfied they do a reasonable job already without fitting and will later seek to use them in real life situations. Note, however, that this will require high quality data on the spread of variants, in particular in terms of times of initial importation in the different jurisdictions.

The first model (Section 2 and Appendix A) is used mainly to evaluate the baseline responses of the paradigm used in the three models to describe the spread of two variants in a population. From this simplest model, we draw two main conclusions, illustrated respectively by Figures 3 and 4. The first is that it takes quite a while before a novel variant is becomes established to the point that it is detected frequently compared to the resident variant. (Recall that the way we encode detection means that the red curve in the bottom panel of Figure 3 shows the ideal situation where all detected cases are detected at the end of their incubation period; so, in practice, the curve should be rising even more slowly.) Since actual identification of the novel variant involves further tests (sequencing, etc.), this means that it is likely that a novel variant is well established before it is effectively detected. Figure 4 points to one of the issues arising in travel control measures: the likelihood that a novel variant successfully establishes itself in a population is highly dependent on the size of the initial number of individuals bearing this variant introduced in the population. While we do not consider the problem of importation size in our investigation of later models, this is indicative of the importance of ensuring that travel is safe. Indeed, consider the case where a location would receive a single transport conveyance before it completely shut down travel. In the conditions of Figure 4, if this conveyance contained a single infected individual, then the probability of a successful importation would be about 1/3. On the other hand, if, while aboard the conveyance, transmission took place and the location received 4 infected individuals, then the importation would be successful in over 80% of the cases.

The second model (Section 3 and Appendix B) focuses on the role of importations. Figure 6 shows the potential strong effect of importations. Note, however, that the curves that show the peak reached within a six months period correspond to unusually high importation rates. To see this, lump together all sources of (direct) importation to the location of interest and assume their total population is *P*^*E*^, that the overall prevalence of variant *X* ∈ *{O, N}* is 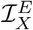, with a fraction 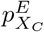 of this prevalence detected. The following formula derived in [5] gives an approximation of the importation rate as 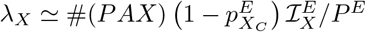, where #(*PAX*) is the number of passengers incoming into the location of interest per unit time. Consider for instance the case *λ* = 2 and assume that, say, *P*^*E*^ = 100, 000 and 75% of cases are detected. Then, we get #(*PAX*) ℐ_*X*_ = 800,000. This means that if the combined prevalence in source locations is, say, 0.5%, then the location of interest is seeing 1,600 incoming passengers per day from the source locations, i.e., inflow of 1.6% its population daily. Recall that in Section 4.3, we computed the pandemic-period inflow into Canada as representing 0.08% of its population per day. Entries into countries typically scale down because of higher self-sufficiency at higher geographic level, but not by two orders of magnitude. Other aspects we consider with the second model are the role of quarantine and of complete travel interruptions. Quarantine is found to be an efficacious way to limit the effects of variant importation; see Figure 9. On the other hand, total interruption of travel is found to be effective only when it is used in the very early stages of propagation of the variant, i.e, within at most 20 to 30 days of the start of potential importations; see Figures 10 and 11. Initial detection of a novel variant may take several weeks or even months in countries carrying out little sequencing of test results. On the other hand, exportation of a novel variant may start very soon after its emergence. From this, it follows that in most cases, total travel interruption is not an efficacious method to fight variant introductions. Further to this, one should bear in mind that there is a high degree of network plasticity, for instance in the air transportation network: cutting an edge typically leads to a reconfiguration of itineraries between points rather than to a substantial attrition of volumes. On the positive side, though, travel interruptions are often implemented in locations that are already implementing quarantine and as such, are subject to lowered rates of importation.

To better understand the dynamics of importations, we consider a third model (Section 4 and Appendix C), written as a simple metapopulation with an exporter and an importer patch connected by movement that is frequency-dependent rather than fixed as in the previous model. A limited preliminary numerical investigation of this complex model shows some interesting characteristics, summarised in Figure 14. The first, seen on both the top and middle row in that figure, is the (entirely expected) existence of a lag between activity in the exporter patch and its “translation” into the importer patch. This lag is (still not unexpectedly) a decreasing function of movement rates. The curve of the percentage of infections resulting from importations in the lower row of Figure 14 is less obvious to interpret. For mobility rates of the order experienced these days (left column), it remains relatively low, representing no more than one infection out of 20 in the left column. Also, it culminates before infection peaks in the exporter patch. For higher movement rates consistent with pre-pandemic travel (right column), importations at times contribute to about 15% of infections, this time with a marked advance on the peak of infection in the exporter patch.

During crises of all types, governments all over the world have also often resorted to a *repli sur soi* : the peril comes from the rest of the world, not from what is done to address the crisis of the day within one’s borders. As noted by [14], “a powerful expression of state’s sovereignty, immigration control provides a typical avenue for governments to reassure their citizens and bolster a national sense of belonging, while providing an ideal scapegoat for their own failure or negligence.” As a consequence, when faced with public health emergencies, travel and border control measures like those discussed here but also of other types (e.g., screening) have been used times and times again. In terms of disease importations, it is indeed true that danger *originally* does come from the rest of the world. However, as can be seen here, a given jurisdiction’s status can very quickly change from being free of infection with a variant to sustaining local transmissions in a context where importations represent but a fraction of the origin of transmission chains and where the jurisdiction itself becomes an exporter.

## Data Availability

The work involves no data. R code may be posted on github at a later date.

## Acknowledgements

JA and SP are supported in part by NSERC Discovery Grants and through the Emerging Infectious Disease Modelling consortium. JA is supported in part by CIHR through the Fields Institute Mathematical Modelling of COVID-19 Task Force and acknowledges past support both financial and logistical from the Public Health Agency of Canada. PYB acknowledges support form ISCD (Institut des Sciences du Calcul et de la Donnée) and the EU grant MOOD (H2020-874850).

## Appendix A. The base ODE model for two variants

### Appendix A.1. Model equations

The ODE model takes the following form. First, the equation for the evolution of *S* is given by

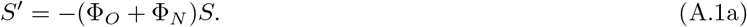

Then, for variant *X* ∈ *{O, N}*,

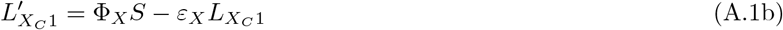

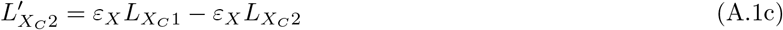

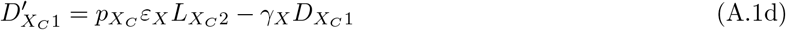

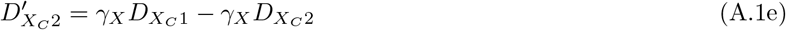

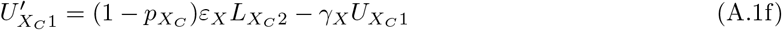

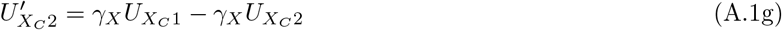

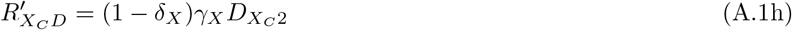

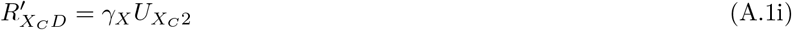

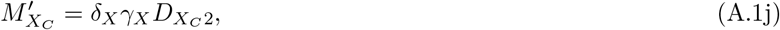

with force of infection

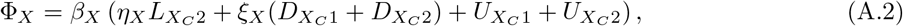

where *η*_*X*_ and *ξ*_*X*_ are the attenuation factors for transmission by incubating and detected cases. Note that the coupling between the resident and new variants dynamics only occurs in the equation for *S*, depicting an implicit competition between the two variants through their infectiousness. This ODE model, as well as the ones following, is considered with nonnegative initial conditions such that at least one of the infected state variables is positive in order to avoid trivial solutions.

#### Appendix A.2. Reproduction number for a single variant

We use the method of [7]. Let us consider variant *X* ∈*{O, N}* in isolation, i.e., in the absence of the other variant. First, we identify the infected variables as

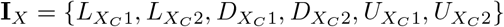

and consider them in that order. Then, using the notation in [7], **D** = 1 since *S* ∈ℝ^*m*^ with *m* = 1. The matrix **Π**_*X*_ is a vector, **Π**_*X*_ = (1, 0, 0, 0, 0, 0)^*T*^, since all new infections move to the 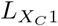 compartment. The row vector **b**_*X*_ describes relative horizontal transmissions and takes the form **b**_*X*_ = (0, *η*_*X*_, *ξ*_*X*_, *ξ*_*X*_, 1, 1). The function denoted *β*(*x, y, z*) in [7] is the constant *β*_*X*_ here. Finally, the matrix **V**_*X*_ describing transitions between and out of infected states for variant *X* takes the form

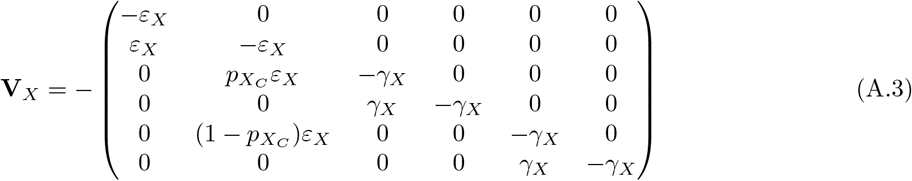

and therefore has inverse

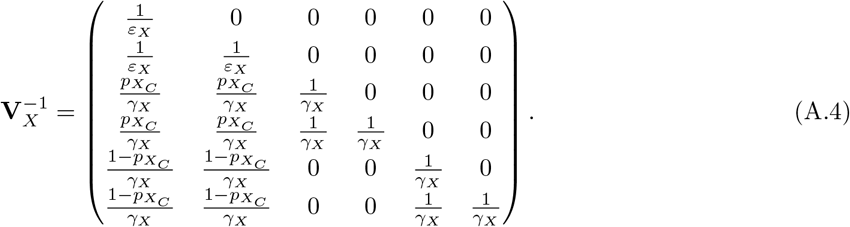

From [7], the basic reproduction number for variant *X* ∈ *{O, N}* is

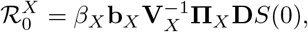

where *S*(0) is the susceptible population at the initial time. Thus, the basic reproduction number for variant *X* ∈ *{O, N}* in the absence of another variant takes the form given by (1).

#### Appendix A.3. Final size for the single variant model

With the notation and considerations in Appendix A.2, we can also use [7] to derive a final size relation for the single variant cases, finding a result almost similar to that found in [9].

#### Appendix A.4. Reproduction number for the two-variant model

The method in [7] cannot be applied to the two variant case because the multigroup nature of the problem leads to a vector incidence function rather than a scalar one. As a consequence, in order to compute the reproduction number for the two-variant system, we use the method of [33], which is easily applied from the considerations in Appendix A.2. With infected variables listed and notation as in Appendix A.2, the matrix **F** used in [33] takes the form

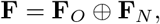

where, for *X* ∈*{O, N}*, **F**_*X*_ has first row given by *β*_*X*_ **b**_*X*_*S*(0) and all other rows zero. For *X* ∈ *{O, N}*, **V**_*X*_ takes the same form as **V**_*X*_ in Appendix A.2. It follows that

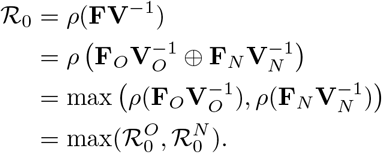

Note that this is as far as the application of [33] can proceed; in particular, it is not possible to apply their Theorem 2 to this epidemic (as compared to endemic) situation. Indeed, there is no situation in the present model where the DFE is unstable; also, the DFE is always locally stable but never locally asymptotically stable.

Intuitively, the issue of final sizes is harder in this setting, as the following thought experiment confirms. Suppose that the two variants each have a value of 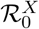 such that the final size for that variant in isolation is larger than *S*(0)*/*2. Clearly, the final size for the entire system is not larger than *S*(0), so final sizes are not additive. However, they cannot operate as the max either, as ℛ_0_ does, since one can expect to see more cases than the maximum implied by either of the final sizes. Refer to, e.g., [3, 20, 21] for more considerations and some results concerning final sizes in multi-group models.

## Appendix B. The ODE importer-only model

### Appendix B.1. Model equations

The ODE model takes the form

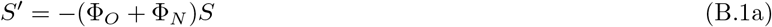

and, for variant *X* ∈*{O, N}*, assuming no importation of detected cases, dynamics in the importation layer are governed by

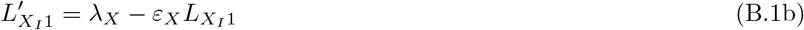

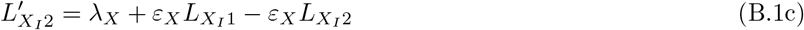

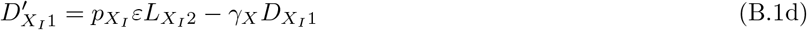

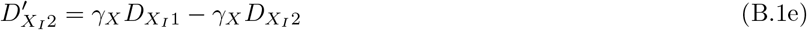

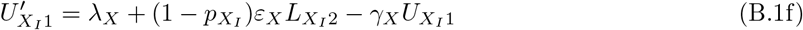

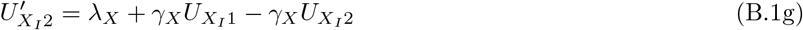

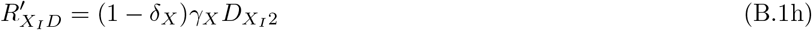

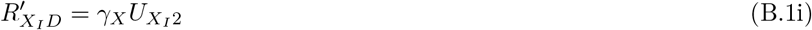

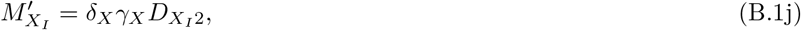

while dynamics in the community are governed by

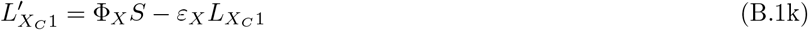

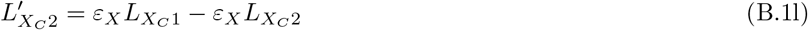

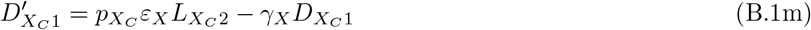

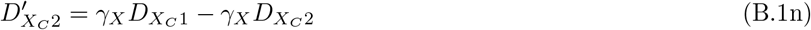

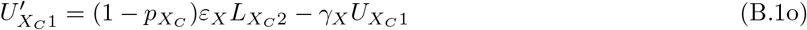

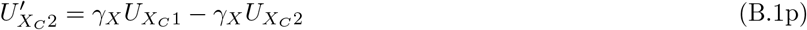

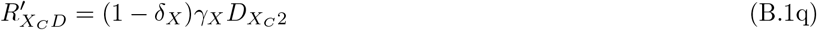

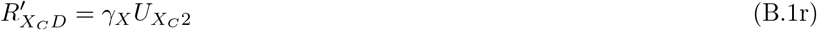

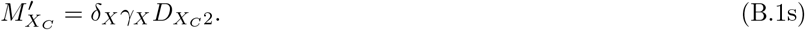

The force of infection takes the form

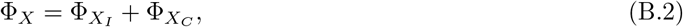

where, for *X* ∈ *{O, N}* and *Z* ∈ *{I, C}*,

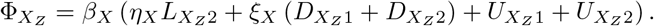

Because of the term *λ*_*X*_ in equations (B.1b), (B.1c), (B.1f) and (B.1g), there is no disease-free equilibrium in the importation layer and as a consequence, it is impossible to compute a reproduction number there, nor for the whole importer-only system.

A reproduction number can however be computed in the community in the absence of connection with the importation layer, giving the same expression as in Appendix A.2. We also use this reproduction number to approximate the value of *β*_*X*_ in the importation layer in the absence of importation. Indeed, as disease propagation parameters are the same, for variant *X* ∈*{O, N}*, in the importation layer and in the community and that contacts from individuals in the importation layer are with susceptible individuals in the community, the *S*(0) used in the computation of 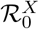 in Appendix A.2 is that of the community. This is the reason why, in (B.2), we use the same *β*_*X*_ for both the importation layer and the community.

### Appendix B.2. Quarantine efficacy

To compute quarantine efficacy, we proceed as in [5]. Suppose an individual arrives in the importation layer while in one of the *unobserved* infected states for variant *X* ∈ *{O, N}*,

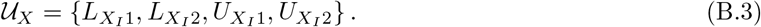

The individual remains in quarantine for *t*_*q*_ time units, during which time they have no contact with anyone else (*β*_*X*_ = 0) and progress through disease stages following the model in the importation layer. If, at the end of the quarantine period, when they start making contacts in the community, they are still in a compartment in *𝒰*_*X*_, then quarantine has *failed* since they can still infect individuals in the community. Quarantine efficacy is thus the complement of the probability that an imported case in *𝒰*_*X*_ is still, after spending *t*_*q*_ time units in quarantine, in one of the states in 𝒰_*X*_.

Consider the matrix of transition rates between stages of infection, where the last three columns of all zeros are omitted:

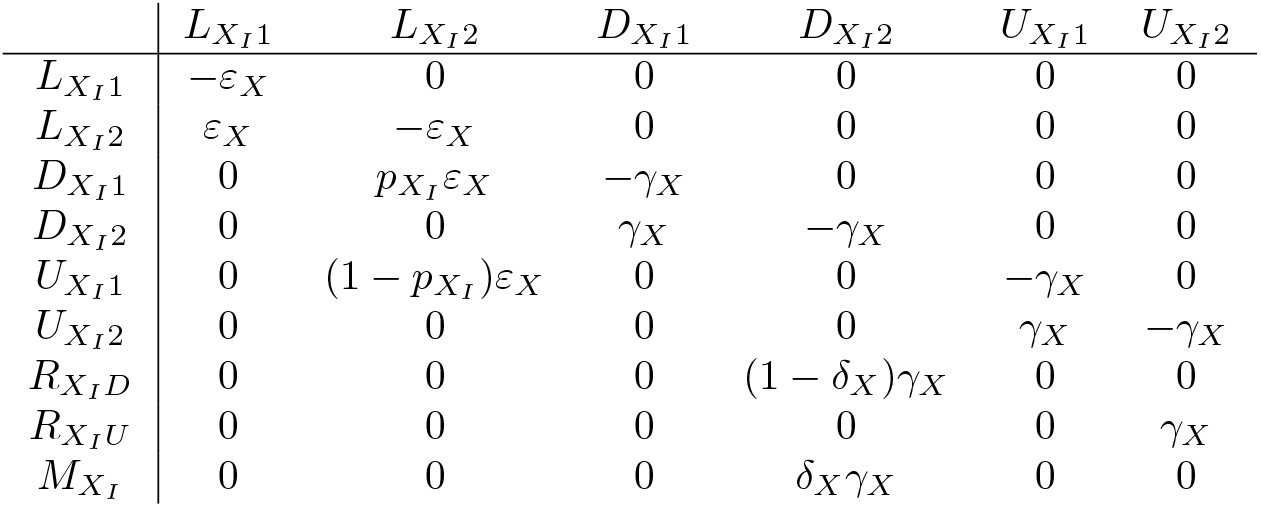

| | $L_{X_I1}$ | $L_{X_I2}$ | $D_{X_I1}$ | $D_{X_I2}$ | $U_{X_I1}$ | $U_{X_I2}$ |
| --- | --- | --- | --- | --- | --- | --- |
| $L_{X_I1}$ | $-\varepsilon_X$ | 0 | 0 | 0 | 0 | 0 |
| $L_{X_I2}$ | $\varepsilon_X$ | $-\varepsilon_X$ | 0 | 0 | 0 | 0 |
| $D_{X_I1}$ | 0 | $p_{X_I}\varepsilon_X$ | $-\gamma_X$ | 0 | 0 | 0 |
| $D_{X_I2}$ | 0 | 0 | $\gamma_X$ | $-\gamma_X$ | 0 | 0 |
| $U_{X_I1}$ | 0 | $(1 - p_{X_I})\varepsilon_X$ | 0 | 0 | $-\gamma_X$ | 0 |
| $U_{X_I2}$ | 0 | 0 | 0 | 0 | $\gamma_X$ | $-\gamma_X$ |
| $R_{X_ID}$ | 0 | 0 | 0 | $(1 - \delta_X)\gamma_X$ | 0 | 0 |
| $R_{X_IU}$ | 0 | 0 | 0 | 0 | 0 | $\gamma_X$ |
| $M_{X_I}$ | 0 | 0 | 0 | $\delta_X\gamma_X$ | 0 | 0 |

Let **T**_*X*_ = [*t*_*ij*_] be the matrix whose entries are given in this table (including the three columns of zeros not shown), with *t*_*ij*_ the rate at which individuals in compartment *j* transition to compartment *i*. Denote *𝒞* (*t*) = (*𝒞*_1_(*t*), …, *𝒞*_*n*_(*t*))^*T*^ the distribution of probabilities of belonging to compartment *i* = 1, …, *n* at time *t*, with 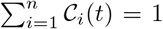 and compartments indexed as in the columns of **T**_*X*_. Then *𝒞* (*t*) satisfies the differential equation

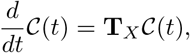

whose solutions are, using the matrix exponential 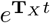,

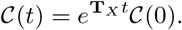

Specifically, entries of 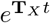 are the probabilities an individual is in the corresponding row-stage at time *t* conditional on having started in the corresponding column-stage at time 0.

If infected individuals with states distributed according to 𝒞 (0) are placed in quarantine for *t*_*q*_ time units starting at time *t* = 0, then the states at time *t*_*q*_ have distribution

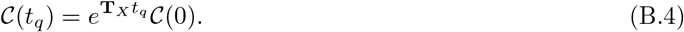

Further, the probability that individuals are in one of the compartments in *𝒰* _*X*_ at time *t ≥* 0 is given by

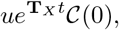

where *u* = (1, 1, 0, 0, 1, 1, 0, 0, 0) is the characteristic vector for undetected infections.

If, after *t*_*q*_ days, the individual is still in an unobservable state 𝒰 _*X*_, then quarantine has failed. Otherwise, quarantine is a success. (Recall that, in the model, *D* and *M* individuals have been detected by authorities, explaining why 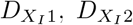 and 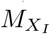 individuals are considered a success of quarantine.) We therefore define the efficacy *c*_*X*_ of quarantine for variant *X* ∈ *{O, N}* as the probability

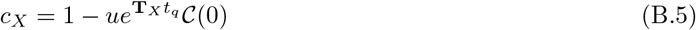

that the imported case is either in an observable state (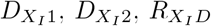 or 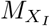) or in the unobserved but recovered state 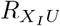. Remark that *c*_*X*_ depends on model parameters through **T**_*X*_ but also on the distribution 𝒞 (0). Typically, it is assumed that 𝒞 (0) = (0.25, 0.25, 0, 0, 0.25, 0.25, 0, 0, 0)^*T*^, i.e., importation of all four unobservable states is equally likely.

Finally, one easily verifies (see [5] for details) that the quarantine-regulated importation rate 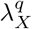 is expressed as

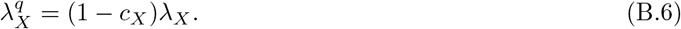

## Appendix C. The ODE exporter-importer model

### Appendix C.1. Model equations

In the exporter, the ODE model takes the following form. First, the dynamics of *S*^*E*^ follows

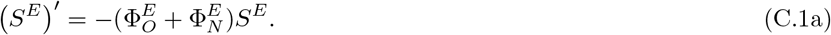

Then, for variant *X* ∈ *{O, N}* and assuming no exportation of detected cases, the dynamics are governed by

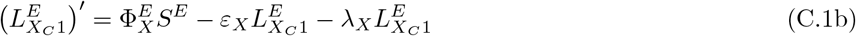

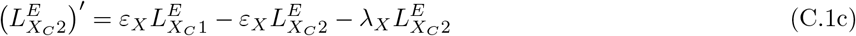

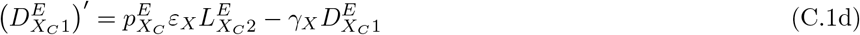

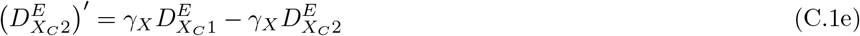

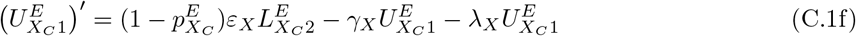

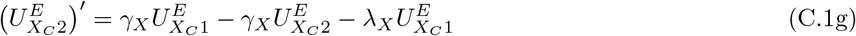

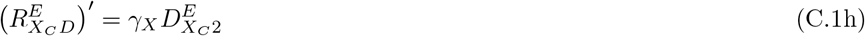

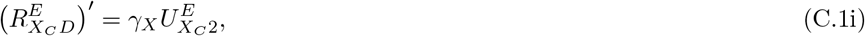

where the force of infection takes the form

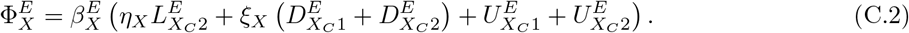

In the importer,

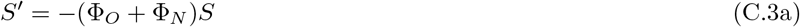

and, for variant *X* ∈*{O, N}* and assuming no importation of detected cases, dynamics in the importation layer are governed by

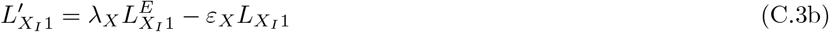

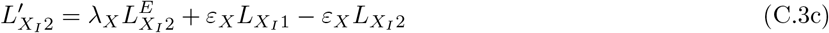

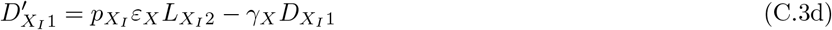

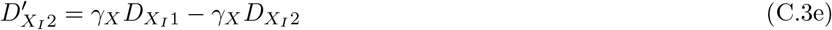

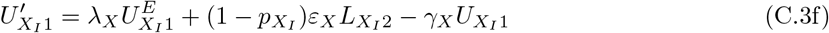

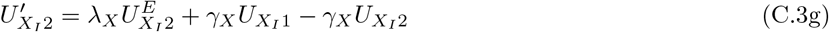

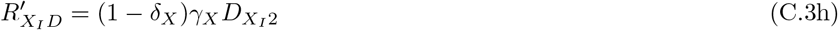

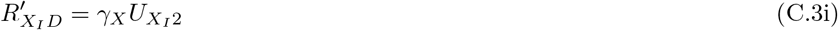

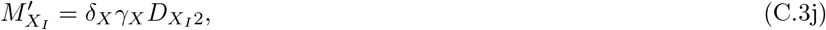

while dynamics in the community are governed by

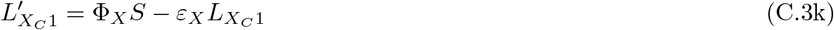

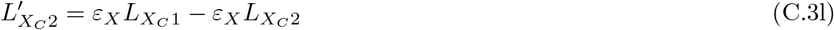

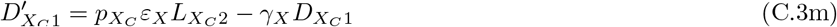

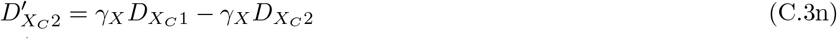

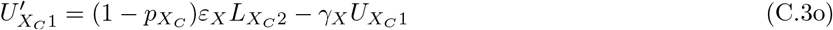

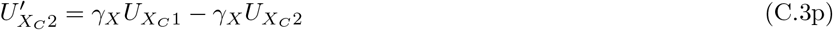

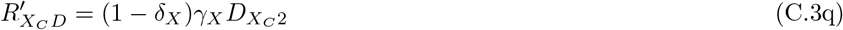

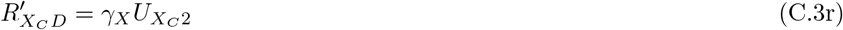

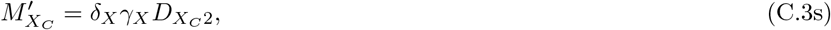

with force of infection

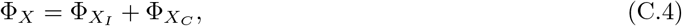

Where, for *X* ∈ *{O, N}* and *Z* ∈ *{I, C}*,

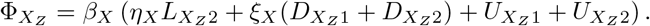

